# How subject-specific biomechanics influences tendon strains in Achilles tendinopathy patients: A finite element study

**DOI:** 10.1101/2024.04.05.24305385

**Authors:** Alessia Funaro, Vickie Shim, Ine Mylle, Benedicte Vanwanseele

**Author notes:** **Correspondence**: Alessia Funaro.

## Abstract

The treatment of Achilles tendinopathy is challenging, as 40% of patients do not respond to the existing rehabilitation protocols. These rehabilitation protocols do not consider the individual differences in the Achilles tendon (AT) characteristics, which are crucial in creating the optimal strain environment that promotes healing. While previous research suggests an optimal strain for AT regeneration (6% tendon strains), it is still unclear if the current rehabilitation protocols meet this condition. Consequently, this study aimed to investigate the influence of a selection of rehabilitation exercises on strains in patients with Achilles tendinopathy using subject-specific finite element (FE) models of the free AT. Secondly, the study aimed to explain the influence of muscle forces and material properties on the AT strains. The 21 FE models of the AT included the following subject-specific features: geometry estimated from 3D freehand ultrasound images, Elastic modulus estimated from the experimental stress-strain curve, and muscle forces estimated using a combination of 3D motion capture and musculoskeletal modelling. These models were used to determine tendon strain magnitudes and distribution patterns in the mid-portion of the AT. The generalized ranking suggested a progression of exercises to gradually increase the strains in the mid-portion of the AT, starting from the concentric and eccentric exercises and going to more functional exercises, which impose a higher load on the AT: bilateral heel rise (0.031 ± 0.010), bilateral heel drop (0.034 ± 0.009), unilateral heel drop (0.066 ± 0.023), walking (0.069 ± 0.020), unilateral heel drop with flexed knee (0.078 ± 0.023), and bilateral hopping (0.115 ± 0.033). Unilateral heel drop and walking exercises were not significantly different and they both fell within the optimal strain range. However, when examining individual strains, it became evident that there was diversity in exercise rankings among participants, as well as exercises falling within the optimal strain range. Furthermore, the strains were influenced more by the subject-specific muscle forces compared to the material properties. Our study demonstrated the importance of tailored rehabilitation protocols that consider not only individual subject-specific morphological and material characteristics but especially subject-specific muscle forces. These findings make a significant contribution to shape future rehabilitation protocols with a foundation in biomechanics.

## 1. Introduction

Achilles tendinopathy is a multifaceted condition (Nicola Maffulli & Kader, 2002) that can profoundly affect individuals (Visser *et al*., 2021), including both athletes and those with relatively low levels of physical activity (Rolf & Movin, 1997). The degenerative changes usually occur in the mid-portion of the free Achilles tendon (AT) (Maffulli *et al*., 2004), with pain as the main symptom. For this reason, the current rehabilitation protocols aim to reduce aggravating loads by introducing pain-relieving loads (Cook & Purdam, 2014). At present, the rehabilitation protocols tend to include a variety of exercises, namely: eccentric exercises (Alfredson et al., 1998a), a combination of eccentric and concentric exercises (Silbernagel et al., 2001) and a combination of more functional exercises (Mascaró et al., 2018). Although the eccentric muscle training is the preferred choice for decreasing pain in chronic Achilles tendinopathy (Silbernagel et al., 2001), there is still little clinical or mechanistic evidence for isolating the eccentric component (Malliaras et al., 2013). Previous animal studies have demonstrated that a certain loading regime (and consequently tendon strains: 6%) promotes tendon remodeling and the creation of a proper (internal) mechanical environment, and therefore is able to reverse early-stage pathological changes in Achilles tendinopathy patients (T. Wang et al., 2015). Also in humans, it was found that a specific threshold of strain magnitude (∼5%) must be reached in order to initiate adaptive changes in both the mechanical and morphological characteristics of the AT (Arampatzis et al., 2007). However, it remains unknown if current rehabilitation exercises provide this optimal strain dose needed for tendon adaptation and regeneration. Indeed, Achilles tendinopathy treatment is still challenging and the failure rate of the classical treatment schemes is still high, with 40% of the patients not responding to these types of training (Nicola Maffulli et al., 2008).

The morphology and material properties of tendinopathic tendons are influenced by changes in the tissue structure, such as the disruption of collagen fiber structure and arrangement and increased type III collagen content (De Mos et al., 2007; J. H. C. Wang, 2006). Indeed, previous investigations have already demonstrated that morphological and material changes are observable in tendinopathic patients, including increased cross-sectional area (CSA) and reduced stiffness and Young’s modulus, in comparison to healthy counterparts (Arya & Kulig, 2010). In the aforementioned study, experimental data of tendon force and elongation were used for the estimation of the tendon strain. However, this experimental measure does not take into account the complex subject-specific geometry of the AT, which makes it unsuitable for the investigation of the internal tendon strains. The AT is the junction of three independent tendons arising from the triceps surae muscles (from soleus (SOL) and gastrocnemius medialis (GM) and lateralis (GL)). Hence, three sub-tendons (SOL, GM and GL, respectively) are fused together in the AT, following the typical twisting structure to the AT. Shim *et al*. (2018) demonstrated that the presence of twist enables a more uniform distribution of stress across the AT, when subjected to differential forces from the triceps surae muscles. Different twisted morphologies were observed among individuals (Edama *et al*., 2015), with the least twisted configuration being predominant (Pękala *et al*., 2017). Hence, it is essential to account for this twisted sub-tendon geometry to better understand the distribution of stress (and strain) within the AT under load, which will reflect the key morphological features observed in individuals. Moreover, the differential force exerted by the triceps surae muscles plays a crucial role in the elongations of the sub-tendons, subsequently the strains in the AT. In individuals with Achilles tendinopathy, there is a noticeable change in the force distribution strategies employed by the triceps surae muscles during dynamic tasks (Mylle et al., 2023). Consequently, it becomes pivotal to consider the impact of triceps surae muscle forces on AT when examining the factors contributing to strain development in patients with Achilles tendinopathy.

Our previous study (Funaro et al., 2022) showed that, finite element (FE) models of the AT can be used to estimate internal tendon strains during rehabilitation exercises, especially under the influence of varying tendon geometry and material properties. Yin *et al*. (2021) showed how different AT morphologies experience different stress and strain distributions, implying that the injury risk is varying between individuals. Moreover, Hansen *et al*. (2017) and Shim *et al*. (2014) pointed out that the free AT stress is highly dependent on subject-specific tendon geometry. Also each individual possesses unique material properties (Yin et al., 2021). These combined findings suggest that personalized geometry and material properties are essential features that should be included in AT FE models when investigating tendon strains during dynamic exercises, especially in patients with Achilles tendinopathy. Previous AT FE studies that incorporated subject-specific geometry and material properties mainly used data from healthy individuals, which do not account for the altered AT geometry and material properties, typical of Achilles tendinopathy patients. When altered AT material and geometry in Achilles tendinopathy patients were considered (Shim et al., 2019), the number of developed FE models was limited (n=8) and did not include the sub-tendon structures. Additionally, muscle forces boundary conditions were recorded during a maximum voluntary isometric contraction, and not during dynamic conditions, such as rehabilitation exercises. Devaprakash *et al*. (2022) demonstrated that estimates of AT strain vary significantly depending on the specific rehabilitation tasks undertaken. Nevertheless, the strain in the AT was determined in a healthy cohort and using external measures that did not consider the tendon’s geometry or material properties. Consequently, these external measures are inadequate to describe the internal load on the tendon, which is more suited for FE analysis. In fact, little is known about the influence that subject-specific muscle forces have on AT strains, in Achilles tendinopathy patients.

The main goal of this study was to evaluate how different rehabilitation exercises affect tendon strains in individuals with Achilles tendinopathy, with the aim of ranking them according to the average strain observed in the mid-portion of the AT. Moreover, we aim to get better insights into the individual differences in tendon strains during different rehabilitation exercises in patients with Achilles tendinopathy and to explain the influence of muscle forces and material properties on the AT strain. To do this, we developed subject-specific FE models of 21 Achilles tendinopathy patients, which includes subject-specific geometry, material properties and muscle forces during commonly used rehabilitation exercises. Specifically, we included exercises which are part of the eccentric muscle strength training regimen advocated by Stanish *et al*. (1986), due to its promising outcomes in the management of Achilles tendinopathy (Niesen-Vertommen *et al*., 1992). We also included concentric exercises, as recommended by Silbernagel *et al*. (2001), for comparative analysis with eccentric exercises. Moreover, we included exercises to compare isolated vs. functional and exercises variation with differential muscle forces. We hypothesized that the strains are lower in concentric and eccentric exercises and gradually increase in more functional exercises in patients with Achilles tendinopathy. We also expect that subject-specific strains are more closely related to the subject-specific muscle forces than the material properties.

## 2. Materials and Methods

### 2.1 Participants characteristics

Twenty-one participants with mid-portion Achilles tendinopathy (17 males, 4 females; age: 49 ± 13 years, height: 178 ± 8 cm, weight: 75 ± 12 kg, average ± SD) participated in the study. The participants met the following inclusion criteria: I) history of intermittent episodes of AT pain lasting more than 6 consecutive weeks within the past 5 years; II) more than one episode of tendon pain exacerbation and remission within the past 5 years; III) palpable focal thickening of the AT in the mid-substance; IV) pain originating from the AT on palpation of thickened AT; and V) sonographic evidence of tendinopathy. Individuals with Achilles tendinopathy were excluded from the study if they reported any of the following: I) history of previous surgery or tears involving the AT; II) systemic diseases affecting collagenous tissue; III) insertional Achilles tendinopathy, calcaneal spurs, plantar fasciitis, and other conditions affecting the foot and ankle complex. All participants completed the Victorian Institute of Sport Assessment-Achilles questionnaire (VISA-A) (Craig et al., 2003), which provides an index of Achilles tendinopathy pain and function (VISA-A score: 73 ± 19). The study was approved by the Ethical Committee UZ/KU Leuven and all relevant ethical guidelines, including provision of a written informed consent prior to participation in the study, were followed.

### 2.2 Experimental data

#### 2.2.1 Data collection

The data collection was performed on patients with Achilles tendinopathy, after a clinical examination and before starting the rehabilitation program, so that all participants were evaluated at the same stage. Upon arrival in the laboratory, the participants were asked to lie prone on an isokinetic dynamometer (Biodex Medical Systems, Shirley, New York, USA) with their knee and hip fully extended, with a fixated foot and a neutral ankle angle. The protocol started with a familiarization of the plantarflexion task and a standardized warm up. Each participant performed two repetitions of a 5s maximum voluntary isometric contraction (MVC). A rest period of 120 s was allowed between contractions. The MVC was repeated if the difference between the contractions was > 8%. After applying a 150ms moving average, the contraction with the highest torque on the tendinopathic side was chosen to determine the target torque (30% and 60% MVC) in the subsequent testing task.

During this second task, 3D free-hand ultrasound (3DfUS) images were acquired during resting condition and isometric contractions at 30% and 60% MVC. A conventional 2D ultrasound machine with a 40-mm linear transducer (L15-7H40-A5, ArtUs EXT-1H system, UAB Telemed, Vilnius, Lithuania) was used to record images of the AT. The ultrasound machine was combined with an optical motion tracking system (V120:Trio tracking system, Optitrack, Corvallis, OR, USA) to generate a 3D reconstruction of the AT. First, the AT of the participants was scanned during resting condition. These images were used for the generation of the subject-specific mesh. Then, participants were asked to practice holding isometric contractions at 30% and 60% MVC until they became familiarized with the testing procedure. Afterwards, the isometric contractions testing task could start. In total, participants performed six contractions to reach a torque of 30% and 60% of MVC (three contractions for each condition, randomized order). Fatigue was monitored by keeping 120 s resting time between contractions. Pain was monitored by asking the participant to give a pain score, between 1 and 10. Extra resting time was given if needed. Feedback of the torque was provided using visual feedback displayed on a monitor in front of the participant.

Subsequently, the linear probe was placed longitudinally over the AT to acquire a static ultrasound image at rest, with the center of the probe aligned with the center of rotation (COR), which is the inferior tip of the lateral malleolus of the tendinopathic ankle. The distance from the inferior tip of the US probe to the COR was measured using a tape measure (*d_1_*) (Merza et al., 2021). This measure was used later for the estimation of the moment arm, as described in the following section. The moment arm was necessary to calculate the force value from the MVC, used for the estimation of the Elastic modulus.

#### 2.2.2 Estimation of subject-specific Elastic modulus

The subject-specific Elastic modulus was estimated using the 3DfUS experimental data. All 3DfUS images were segmented in the 3D Slicer (Version 4.11.20210226) software. To determine subject-specific tendon length, two main anatomical landmarks (calcaneal notch and soleus muscle-tendon junction) were manually located on 2D images of the AT and the tendon length was defined as the point-to-point distance between these two landmarks. Tendon elongation was calculated for both 30% and 60% MVC, by subtracting tendon lengths in the two contractions from the corresponding resting length, which was used to calculate the overall tissue strain for each subject by dividing the subject’s tendon elongation by the subject’s resting length. Then, the subject-specific tendon force was calculated by dividing the subject-specific MVC by the subject-specific moment arm of the AT. The subject-specific moment arm was estimated from 2D images. First, the distance between the skin and midline of AT (*d_2_*), also known as the line of action (Maganaris *et al*., 1998), was measured. Then, the difference between *d_1_* (explained in data collection section) and *d_2_* represented the AT moment arm (Merza et al., 2021). Tendon cross-sections were digitized manually from the 3D images at half distance between the calcaneal notch to the soleus muscle-tendon junction, to estimate the subject-specific average CSA of the mid-portion of the AT. CSA was estimated at rest, 30% and 60% MVC. Subject-specific tendon stress was then obtained by dividing the calculated subject-specific tendon force with the subject-specific CSA of the tendon. Finally, the subject-specific Elastic modulus was calculated as the slope of the line fitted to the subject-specific stress-strain values at 30% and 60% of the peak force for each subject. The subject-specific values of the Elastic modulus can be found in the **Supplementary Material**. The average values and the standard deviations of MVC, CSA at rest and Elastic modulus among the 21 participants can be found in **Table 1**.

**Table 1.** The table shows the average of the subject-specific values for maximuml voluntary contraction (MVC), moment arm, cross-sectional area (CSA) and Elastic modulus.

| MVC (Nm) | Moment arm (mm) | CSA (mm<sup>2</sup>) | Elastic modulus (MPa) |
| --- | --- | --- | --- |
| 113 ± 32 | 45 ± 5 | 112 ± 31 | 632 ± 201 |

#### 2.2.3 Estimation of subject-specific muscle forces

The participants came for a second time to the Movement and posture Analysis Laboratory Leuven (Belgium) to complete five repetitions of six rehabilitation exercises in a randomized order: walking (walk), bilateral heel rise (birise), bilateral heel drop with extended knee (bidrop), unilateral heel drop with extended knee (unidrop), unilateral heel drop with flexed knee (unidrop bent) and bilateral hopping (bihop). In between trials, the participant was given a minimum of 30 s of rest before moving on to the next trial. An extended Plug-in Gait marker set including 34 retroreflective markers, of which the trajectories were recorded using ten infrared cameras (Vicon, Oxford Metrics, Oxford, United Kingdom) at a sampling rate of 150 Hz, were placed on anatomical landmarks to obtain kinematic data. Ground reaction force data was measured from the participant’s affected leg using a force plate embedded in the walkway. A modified generic musculoskeletal model (OpenSim gait2392 model) (Delp et al., 1990) with six degrees of freedom and 43 Hill-type muscle-tendon actuators per leg was scaled in OpenSim 3.3 (OpenSim, Stanford, CA, United States) and joint kinematics were then computed using a Kalman Smoothing algorithm (F. De Groote et al., 2008). Next, an inverse dynamic approach was used to calculate the joint moments. The forces of the three triceps surae muscles (SOL, GM, and GL muscle forces) were then estimated using a dynamic optimization method by minimizing the sum of squared muscle activations (Friedl De Groote et al., 2016). The subject-specific muscle forces at the time of peak total muscle force during each exercise were used as boundary conditions in the subject-specific FE models. The subject-specific values of the muscle forces can be found in the **Supplementary Material**.

### 2.3 Generation of FE models

#### 2.3.1 Development of the three types of AT FE models

For each participant, three FE models of the AT were developed. The first model, which we call “*subject-specific model”*, was developed with subject-specific geometry, material properties and muscles forces to determine the individual ranking of the exercises based on AT strains. Subsequently, to investigate the impact that subject-specific material properties and muscle forces have on AT strains, two additional models were developed. The first model, called “*material model*”, had subject-specific Elastic modulus and generic muscle forces. The other model, called “*muscle forces model*”, had subject-specific muscle forces and generic material properties. The generic muscle forces were estimated as average of the subject-specific muscle forces, for each exercise (**Table 2**), among all the participants. The generic Elastic modulus was estimated as the average of the subject-specific Elastic moduli (**Table 1**), among all the participants. All the models had subject-specific geometry. A summary of the characteristics of the three models is presented in **Figure 1**.

**Table 2.** The table presents the generic muscle force values for six rehabilitation exercises, used as boundary conditions for loading the three sub-tendons (SOL, GM, and GL) of the FE models. These generic muscle forces were determined by averaging subject-specific muscle forces from the triceps surae muscles across 21 participants, across the six rehabilitation exercises (in the table: mean ± SD, measured in Newton (N)).

|  | SOL force (N) | GM force (N) | GL force (N) |
| --- | --- | --- | --- |
| **BiRise** | 827.6 ± 184.4 | 205.5 ± 37.9 | 91.0 ± 18.5 |
| **BiDrop** | 847.7 ± 178.6 | 246.6 ± 71.7 | 117.6 ± 28.8 |
| **UniDrop** | 1852.7 ± 671.3 | 439.7 ± 119.4 | 149.7 ± 35.2 |
| **Walk** | 1603.6 ± 402.8 | 627.4 ± 219.9 | 187.7 ± 63.4 |
| **UniDrop Bent** | 2324.4 ± 636.9 | 430.1 ± 141.2 | 145.5 ± 80.6 |
| **BiHop** | 3565.2 ± 680.8 | 416.6 ± 270.5 | 82.02 ± 100.5 |

**Figure 1.**
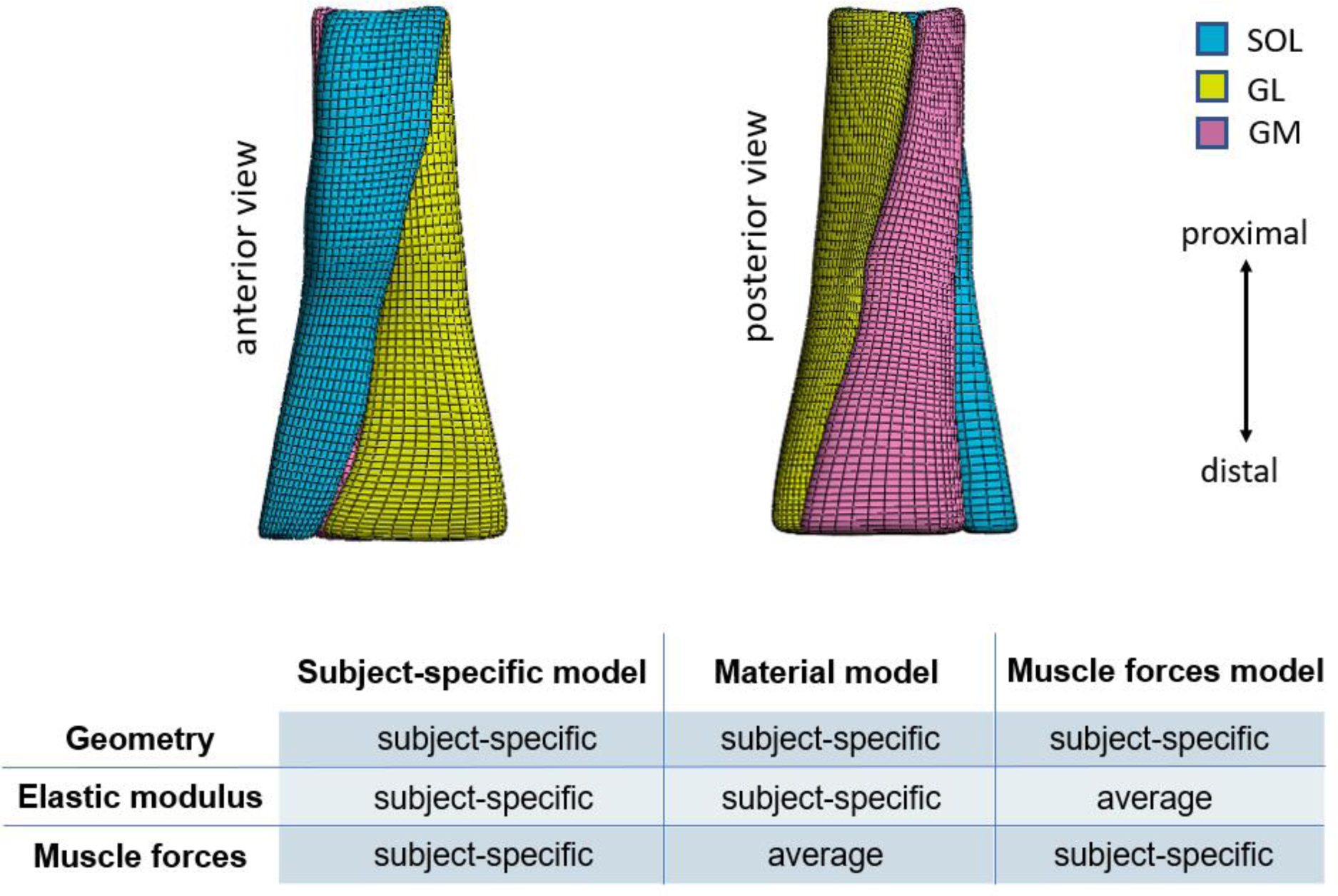
On the top, anterior and posterior view of an example of Achilles free tendon geometry, representing the least twisted geometry described by Pekala et al. (2017). The 3D tendon models were divided in three sub-tendons, each one arising from one of the three triceps surae muscles: the soleus (SOL) and the two heads (medialis, GM and lateralis, GL) of the gastrocnemius muscle. On the bottom, a summary of the characteristics of the three different models.

#### 2.3.2 Generic model geometry

A first generic template mesh was developed, to allow the consistent definition of the three sub-tendons within subject-specific FE models. This mesh was generated from an initial geometry, obtained by segmentation of images from one healthy male subject (age = 22 years, weight = 64 kg, height = 180 cm) recorded by 3DfUS images, defining the outer geometry of the tendon. In this study, only the least twisting geometry described by Pękala *et al*. (2017) was created, since the ranking of the rehabilitation exercises is not influenced by the twist of the sub-tendons (Funaro et al., 2022). This mesh was developed using Materialise 3-matic (Materialise NV, Leuven, Belgium). The tendon model was divided into three sub-tendons and the twisting structure was defined. The sub-tendons geometries were meshed into 8-nodes hexahedral solid elements. A mesh convergence study was performed to refine the mesh until the Principal Effective Lagrange strains reached an asymptote.

#### 2.3.3 Constitutive models

The geometry was represented as an incompressible, transversely isotropic hyperelastic material (Weiss *et al*., 1996), which we already used in our previous study (Funaro et al., 2022). The uncoupled strain energy function can be written as follows:

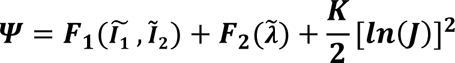

Here, 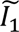 and 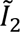 are the first and second invariants of the deviatoric version of the right Cauchy-Green deformation tensor. 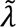 is the deviatoric part of the stretch along the fiber direction, and *J* = *det(F)* is the Jacobian of the deformation. This strain energy density function consists of two parts: *F_1_* represents the material response of the isotropic ground substance matrix, while *F_2_* represents the contribution from the collagen fibers. *F_1_* was described as a Neo-Hookean model and it was equal to 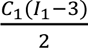. The resulting fiber stress from the fibers was expressed using the following piecewise function:

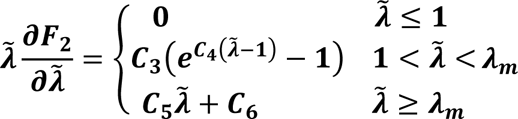

Here, λ_*m*_ is the stretch at which the fibers are straightened, *C_3_* is the scaling of the exponential stress, *C_4_* is the rate of the uncrimping of the fibers and *C_5_* is the Young’s modulus of the straightened fibers. *C_6_* is determined from the requirement that the stress is continuous at λ_*m*_. For the *subject-specific model* and *material model*, each participant had a different *C_5_* value, calculated using the subject-specific stress-strain curve as described above. For *muscle forces model*, *C_5_* value was the average of the subject-specific Elastic moduli (**Table 1**). The values of the other material properties were not subject-specific and they were obtained from Shim *et al*. (2019), involving tendinopathic tendons. The constitutive models implemented in FEBio Studio (Maas et al., 2012) as “*trans iso Mooney-Rivlin*” were used in this study. The sub-tendons fascicles were modelled using the FEBio’s local fiber direction function (*a_0_*) (Maas *et al.,* 2012), which was defined for each element to represent the tendon fascicle structure (Knaus & Blemker, 2021). For each sub-tendon, fibers were directed from the proximal cross-section to the distal cross-section. In this way, the resulted local fibers could follow the twisting structure of the sub-tendons.

#### 2.3.4 Creation of FE models with subject-specific geometry with free-form deformation

Subject-specific FE models were generated using the subject-specific geometry. First, the 2D ultrasound images of the tendon during resting conditions were transformed into the global coordinate system using the 3D Slicer software to create a reconstructed 3D volume. After computing a 3D volume reconstruction, the AT was manually outlined using the reconstructed 2D images, and the corresponding borders were combined to generate 3D data clouds that captures tendon geometry. In total, 21 subject-specific 3D shapes were obtained. Subsequently, the template mesh containing three sub-tendons was customized to each subject-specific geometry obtained with the subject’s 3DfUS image. The free-form deformation method (Fernandez et al., 2018) was used, which morphs an underlying mesh by embedding it inside a host mesh. The external and internal nodes of a given mesh are deformed with the same transformation, to match the subject’s geometry. In this way, it was possible to obtain subject-specific free AT geometries for each participant, representing the least twisting structure described in the literature (Pękala *et al*., 2017).

#### 2.3.5 FE models boundary conditions

The models boundary conditions were defined in FEBio (Maas et al., 2012). The contact between the three sub-tendons was defined as frictionless sliding (Knaus & Blemker, 2021). The distal end of the tendon models was fixed to mimic the attachment of the tendon to the calcaneus. The muscle forces for the different rehabilitation exercises were applied as nodal loads to the proximal faces of each sub-tendon. The nodal displacements were constrained to move only in the distal-proximal direction to mimic the constraints provided by paratenon and fascia cruris that hold the sub-tendons together (Diniz et al., 2022).

### 2.4 Internal tissue strain analysis

Analyses were conducted to quantitatively examine strain distribution patterns during various exercises in the FE models. The average of the maximum principal strain in the mid-portion (defined as the middle third of the AT models) of the *subject-specific models* was analysed to rank the rehabilitation exercises for the Achilles tendinopathy patients (**Figure 2**). The ranking of the exercises is based on the mean of the average strains among all the participants. An optimal strain range was used between 5% and 7% tendon strain based on the findings of Wang *et al*. (2015) while allowing for a margin of error of up to 1% strain. Secondly, we investigated the distribution of the maximum principal strain and identified the location and magnitude of the peak maximum principal strain across the *subject-specific models*. This was done to characterize the overall strain patterns associated with different exercises (**Figure 2**). The average and peak of the maximum principal strain in the mid-portion of the *material models* and *muscle forces models* were analysed for subsequent regression analysis.

**Figure 2.**
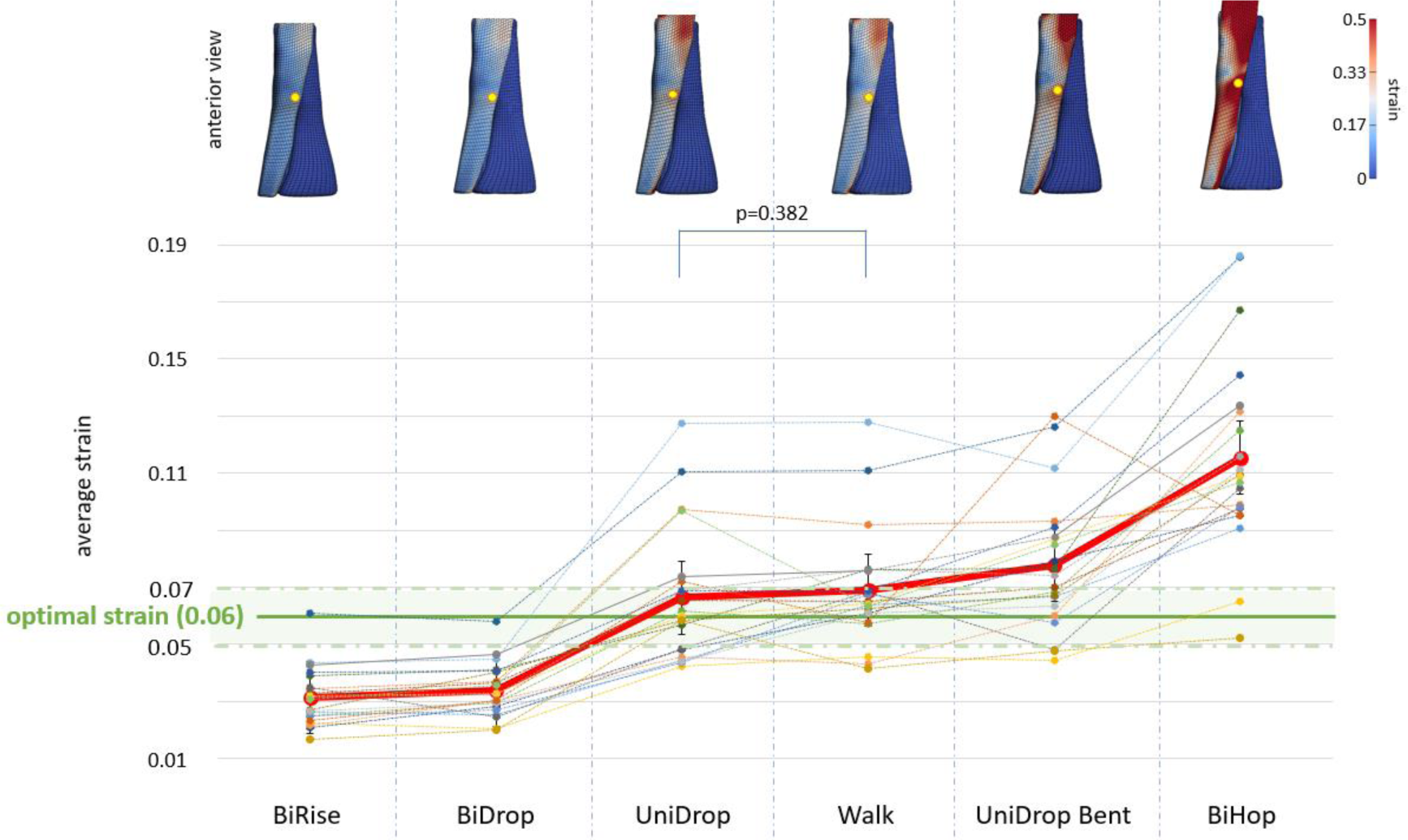
On the top, a representative example of the distribution of the maximum principal strain for each exercise, from one participant. The yellow dots indicate the location of the peak strain. On the bottom, the red line represents the mean of the average strain during each exercise, in the mid-portion of the subject-specific models. Exercises are ranked based on this average strain. The dotted lines represent the average strain during the exercises for each participant, in the mid-portion of the subject-specific models. The optimal strain, designated as 0.06 (6%) according to Wang et al. (2015), is represented by the green bar, which highlights the range defined as 0.06 ± 0.01 (6±1%) strain. All exercises showed statistically significant differences except for unidrop and walk. Significance was set at p<0.05.

### 2.5 Statistical analysis

Statistical analysis for the strains in the mid-portion of the AT models was performed using SPSS Statistics software (IBM Corp. (2017). IBM SPSS Statistics for Windows (Version 29.0.1.0). Armonk, NY: IBM Corp). A repeated measures ANOVA was performed to compare the means of the average and peak strains in the mid-portion of the *subject-specific models* across the exercises. If a significant main effect was found, the LSD (Least Significant Difference) for post-hoc testing was applied to identify the exercises whose means are statistically different. All data are presented as mean ± SD. Statistical level of significance was set as p<0.05. In the subsequent analysis, linear regression was conducted to assess the impact of subject-specific material properties and subject-specific muscle forces on the average and peak strains in the mid-portion of the AT FE models. For significant correlations, the coefficient of correlation (R) between *subject-specific model* and *material model*, and then between *subject-specific model* and *muscle forces model*, was compared.

## 3. Results

### 3.1 Subject-specific models

#### 3.1.1 Ranking of the Rehabilitation Exercises Based on Tendon Strains

Starting from the lowest average strain in the mid-portion of the *subject-specific models*, the generalized ranking of rehabilitation exercises was: birise (0.031 ± 0.010), bidrop (0.034 ± 0.009), unidrop (0.066 ± 0.023), walk (0.069 ± 0.020), unidrop bent (0.078 ± 0.023), and bihop (0.115 ± 0.033). All exercises demonstrated statistically significant differences from one another, except for unidrop and walk, as depicted in **Figure 2**. Notably, at peak total muscle force, only unidrop and walk showed average strains within the range of the reported optimal strain, when the generalized ranking was evaluated. However, when evaluated on an individual level, the variations in average strain across patients become more evident not only in the exercise rankings but also in the exercises that fall within the optimal strain range. This indicates that participants achieved optimal strain levels during different exercises. While the majority of participants reached optimal strain levels during the unidrop and walk exercises (eight and twelve participants, respectively), individual rankings revealed that the optimal strain range could also be attained during the unidrop bent exercise for seven participants, bihop for two participants, birise for one participant, and bidrop for one participant as well (**Figure 2**).

#### 3.1.2 Strain distribution, peak location and peak strain values

The peak strain consistently occurred in the middle of the mid-portion or upper mid-portion of the SOL sub-tendon, which is the sub-tendon exposed to the highest muscle forces (**Table 2**). However, there was marked variability in the exact location of the peak strain and strain distribution among patients, as illustrated in **Figure 3**. Conversely, within a patient, both the peak location and strain distribution remained consistent throughout the exercises (as shown in **Figure 2**). Nevertheless, the magnitude of peak strain varied depending on the load applied during each exercise. The peak strain values in the mid-portion of the subject-specific models were: birise, 0.194 ± 0.081; bidrop, 0.196 ± 0.069; unidrop, 0.481 ± 0.299; walk, 0.403 ± 0.209; unidrop bent, 0.648 ± 0.389; bihop, 0.963 ± 0.462. All exercises exhibited significant differences (p < 0.05) from each other, except for birise and bidrop (p=0.774), and walk and unidrop (p = 0.061).

**Figure 3.**
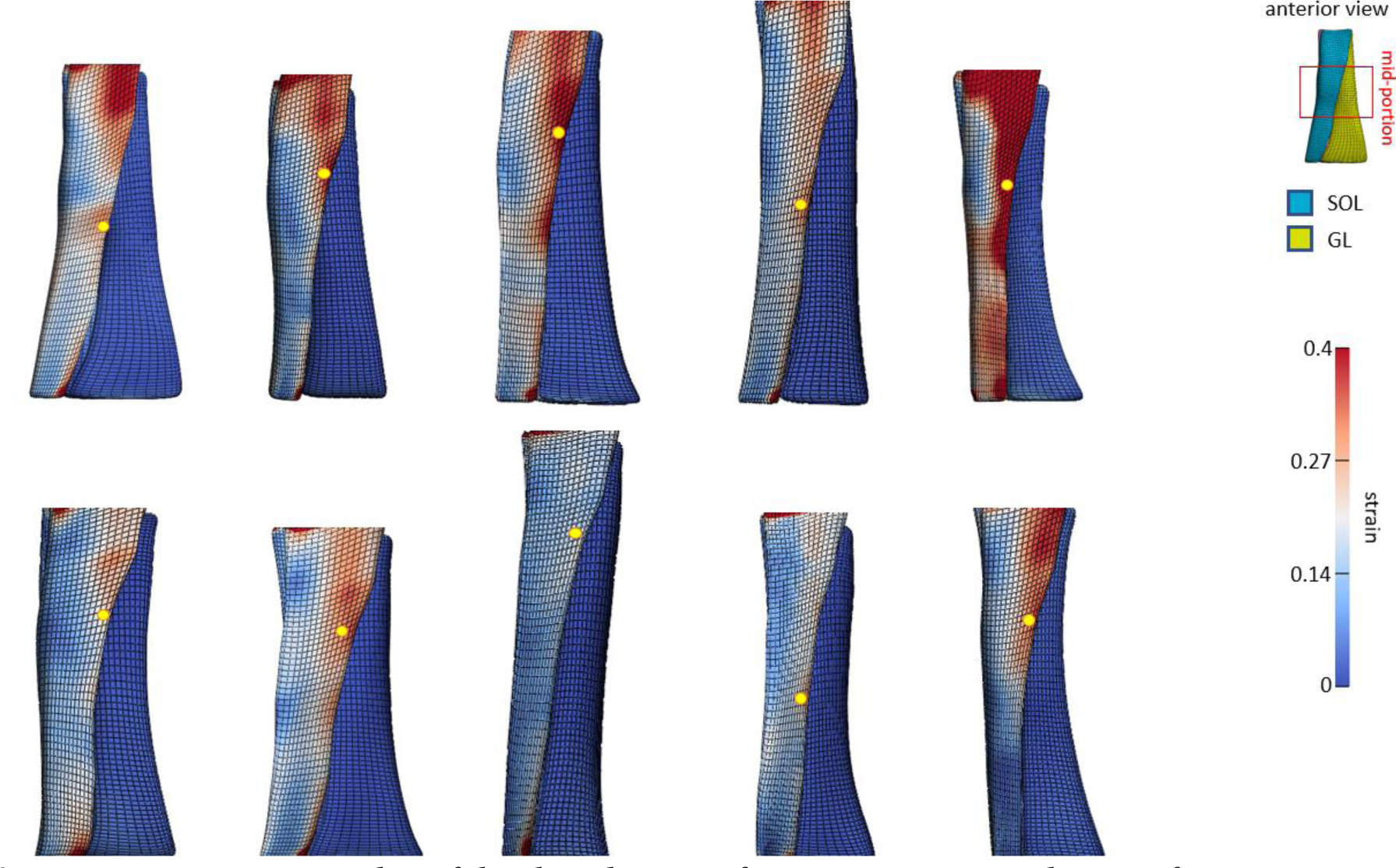
Representative examples of the distribution of maximum principal strain for ten participants during walking. The anterior view of the subject-specific models is presented, highlighting the SOL sub-tendon with the highest strains. The yellow dots indicate peak strains, predominantly situated in the middle and the upper part of the mid-portion of the SOL sub-tendon. The mid-portion refers to the middle third of the model, as shown in the example in the top right of the figure. Location of the peak strain is determined over the whole free tendon, ignoring strains in the regions where boundary conditions were applied, in accordance with Saint-Venant’s principle.

### 3.2 Material models and Muscle forces models

#### 3.2.1 Regression analysis

Scatterplots depicting the average and peak of the maximum principal strain in the mid-portion of the AT models for the *material models* and the *muscle forces models* were presented in **Figure 4**. The association between the *subject-specific model* and the *material model* yielded a lower R than the association between the *subject-specific model* and *muscle forces model*. The higher similarity in strain magnitude between the *muscle forces model* and the *subject-specific model* (as observable in **Figure 5**) indicates that the influence of subject-specific Elastic modulus has a lesser impact on the strains of the *subject-specific model* compared to the influence of muscle forces.

**Figure 4.**
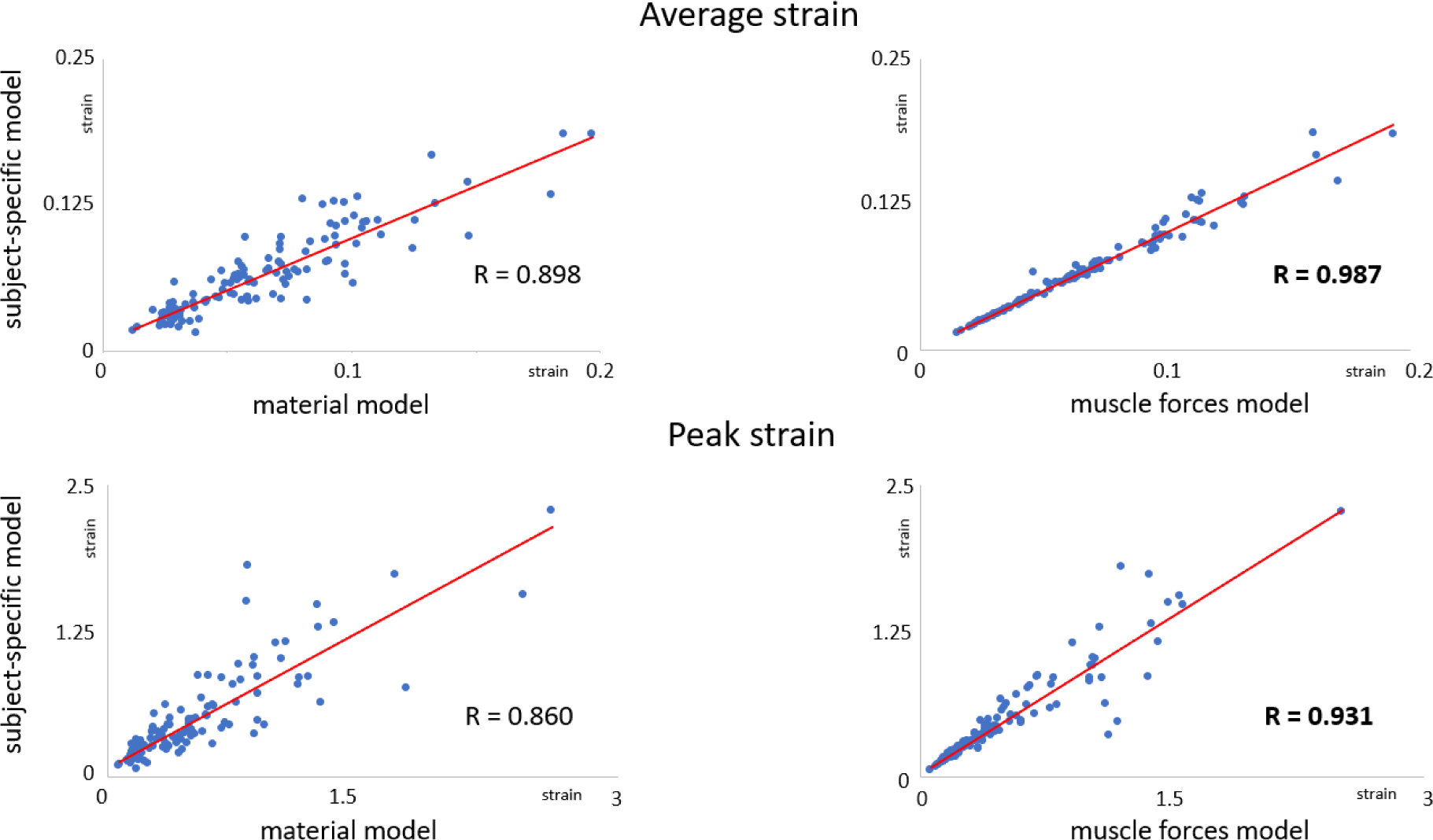
Scatterplots and linear regression lines of comparisons of subject-specific model with material model and muscle forces model, for the average (on the top) and peak (on the bottom) strains in the mid-portion of the AT models. The coefficient of correlation (R) computed across all the participants and exercises is also displayed in each plot.

**Figure 5.**
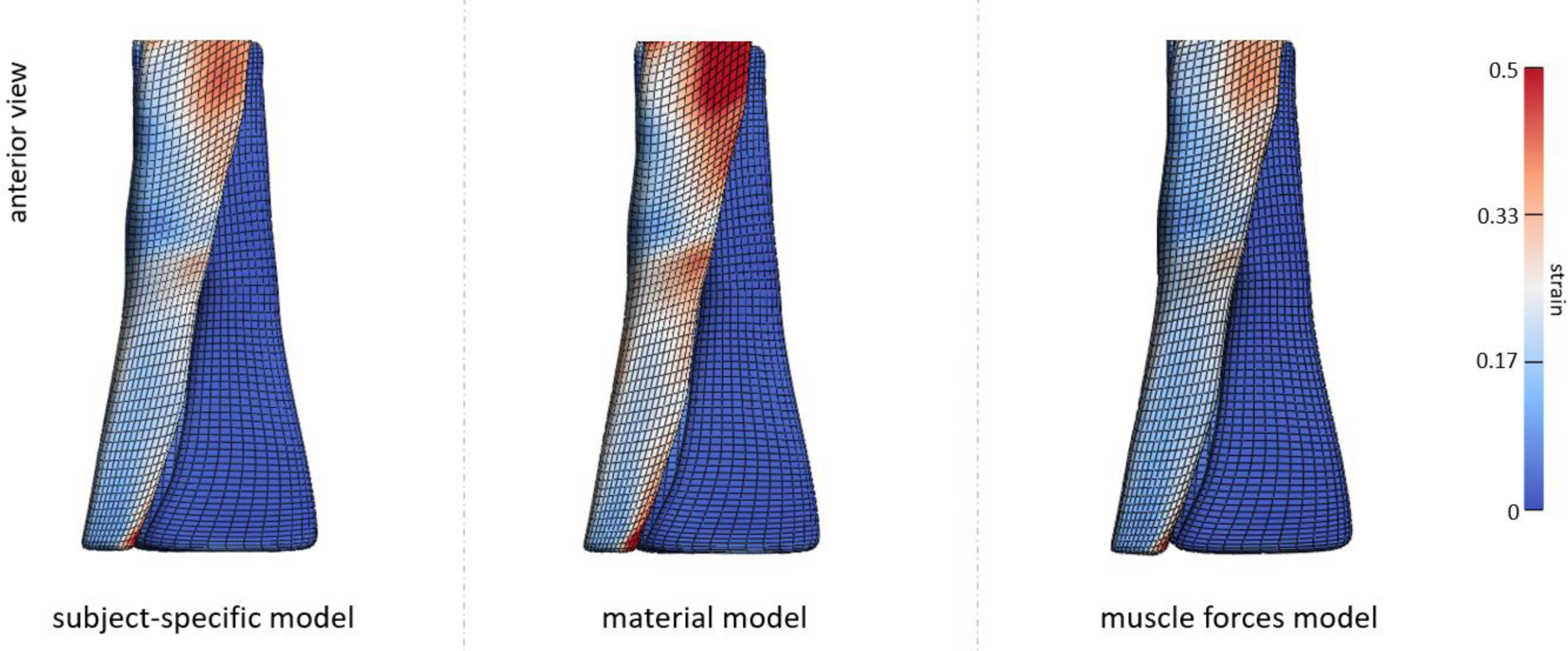
Representative examples of the strain distribution for the three types of AT models developed: subject-specific model, material model and muscle forces model. The example is presented for one participant, during the simulation of walking. While the strain distribution and peak strain location remain constant among models, the image evidences the high correlation of the strain magnitude between the subject-specific model and the muscle forces model, as demonstrated by the high R value.

## 4. Discussion

The primary objective of this research was to assess the tendon strains during various rehabilitation exercises in patients with Achilles tendinopathy, aiming to rank the exercises based on average strain in the mid-portion of the AT. Critical in the estimation of patient-specific strains is the application of FE models incorporating subject-specific geometry, material properties, and muscle forces. This study successfully determined a ranking of the rehabilitation exercises aimed at gradually increasing the strains in the mid-portion of the AT. The recommended progression begins with concentric and eccentric exercises, such as bilateral heel rise, bilateral heel drop and unilateral heel drop, and advances to more functional exercises like walking, unilateral heel drop with flexed knee and bilateral hopping. Both unilateral heel drop and walking exercises demonstrated no significant difference and fell within the optimal strain range. However, when analyzing individual strains, there was variability in exercise rankings among participants, as well as exercises falling within the optimal strain range. Lastly, variability between participants in strains were more closely related to the subject-specific muscle forces rather than the material properties.

### 4.1 Ranking of the Rehabilitation Exercises Based on Tendon Strains

In accordance with our hypothesis, the resultant generalized ranking delineated a sequence of rehabilitation exercises to gradually increase tendon strains, starting with the relatively low-load concentric bilateral heel rise, and progressing towards the eccentric bilateral heel drop. While Alfredson *et al*. (1998) recommended to incorporate the unilateral heel drop exercise during the initial phase, Silbernagel *et al*. (2001) suggested it be introduced in a later stage of the rehabilitation program, due to concerns about tendon overload. Our results demonstrate that the unilateral heel drop induced higher strains in the AT compared to the two bilateral exercises. However, this increased load caused by the unilateral heel drop achieves optimal strain levels (T. Wang et al., 2015) which suggests that it can already be used in the initial phase of the rehabilitation program. Similar to other studies (Devaprakash *et al*., 2022; Franz *et al*., 2015), we confirmed that also walking falls within the optimal strain range necessary for AT tissue healing, despite the fact that walking exposes the AT to a load approximately four times body weight (Komi *et al*., 1992). This suggests that individuals with Achilles tendinopathy may safely engage in walking activities as part of their rehabilitation program from the beginning, without significant disruption to their daily routines.

In adherence to the Alfredson’s protocol (Alfredson et al., 1998b), we also incorporated the unilateral heel drop with flexed knee exercise. This exercise plays a pivotal role in the rehabilitation program by enhancing the activation of the soleus muscle and facilitating a redistribution of muscle forces. However, the increased activation of the soleus led to increased strains, which surpassed the optimal range. It is known that the AT experiences high loads during hopping, equivalent to approximately five times body weight (Komi et al., 1992). Indeed, our simulation of hopping exercises reflected these high loads, resulting in strains surpassing the optimal threshold. Nevertheless, our calculations of tendon strains during hopping surpassed those documented in earlier research. For example, Lichtwark *et al*. (2005) noted peak strains of 8.3% during unilateral hopping, with tendon strains measured at the gastrocnemius medialis myotendinous junction level. The probable explanation for this discrepancies lies in the fact that our strain assessments were based on the free AT, which undergoes notably greater longitudinal strain compared to the aponeurosis (Farris et al., 2013; Obst et al., 2016). Moreover, the strains that we reported are from computational models, where the strains are measured at the element levels. This includes some localised peak strains that are not captured by global strains measured in above mentioned studies. This might have also caused the higher strains that we observed in our study.

Our simulations were based on FE models developed from patients at the beginning of their rehabilitation program. Considering the likelihood of morphological and material properties alterations throughout the rehabilitation process, it becomes pertinent to explore how these modifications could impact AT strains. This analysis could inform the right timing of activities such as unilateral heel drop with flexed knee and hopping within the rehabilitation program. For instance, Silbernagel *et al*. (2015) have suggested incorporating such activities in a later stage with the return-to-sport program. By evaluating the rehabilitation stages and monitoring strain variations together with morphological and material properties changes, we can ensure that Achilles tendinopathy patients undergo a phased and carefully managed recovery process by appropriate loading of the AT.

The unique characteristics in the subject-specific responses were evident when inspecting the tendon strains for the individual Achilles tendinopathy patients. Based on our results, it becomes clear that a generic approach is insufficient since variations in tendon strains among participants for a specific rehabilitation exercise become visible. While unilateral heel drop and walking were the most common exercises among participants (eight and twelve participants, respectively), optimal strains were observed in seven participants during unilateral heel drop with a flexed knee, supporting the theory of eccentric exercises as the preferred treatment for Achilles tendinopathy patients (Alfredson et al., 1998b). Interestingly, bilateral heel rise and bilateral heel drop exercises consistently resulted in suboptimal strain levels across individual rankings, with only one participant achieving the optimal strain for both exercises. Further exploration of these cases could provide further insights in what characteristics determine these individual tendon strains.

Notably, the participant who achieved optimal strain during bilateral heel rise and bilateral heel drop exhibited an Elastic modulus comparable to the average (709.1 vs. 631.9 MPa, individual and average Elastic modulus, respectively), suggesting that material properties alone may not be decisive in this case. Indeed, anthropometric analysis revealed that this participant had a higher weight compared to the average (80 vs 76 kg, individual and average weight, respectively), and also larger muscle forces during the execution of these exercises. Furthermore, analysis of AT morphology highlighted a significantly smaller CSA (the smallest CSA, more specifically) in the mid-portion of the AT model among the participants (66.4 vs 112.3 mm^2^, individual and average CSA, respectively). The combination of these factors led to greater strains during the performance of two generally low-load exercises and so the achievement of the optimal strains earlier during the progression of the rehabilitation exercises. Therefore, the necessity of personalized exercises prescription tailored to individual muscle forces and tendon morphology becomes evident, since the strains seem to be highly dependent.

The bilateral hopping was the exercise imposing the highest load on the AT for all participants and, for this reason, generally located at the end of the rehabilitation exercises progression, given the high strains produced. Nevertheless, for two participants, this exercise led to optimal strains. Again, individual characteristics such as material and morphological properties of the AT, and muscle forces offer insights into this variation. Indeed, one of these two participants exhibited a similar Elastic modulus and CSA compared to another participant (498.2 vs 478.5 MPa and 111.8 vs 99.8 mm^2^, Elastic modulus and CSA for the first and the second participant, respectively). However, while one of the participants only reached the optimal strain level later in the exercise progression, during bilateral hopping, the other achieved it during walking, demonstrating consistently higher strains compared to the former. The answer to the question “where is this difference coming from?” can be found in the force production: the second participant generated higher strains due to greater muscle forces. This closer analysis also demonstrated that when evaluating strains, it’s crucial to consider all subject-specific characteristics. In this case, neglecting subject-specific muscle forces could have introduced errors into the analysis.

These two examples highlight the significant sensitivity of the AT strains to the individual muscle forces and geometry of each subject. This sensitivity is further supported by the findings of the regression analysis. According to our hypothesis, our investigation established that the strains were closer related to the subject-specific muscle forces rather than the material properties. In line with previous results (Hansen et al., 2017; Shim et al., 2019), it becomes evident that when creating subject-specific models, prioritizing not only subject-specific geometry but also subject-specific muscle forces over material properties, is crucial for a more accurate representation of the AT strains, especially when considering Achilles tendinopathy patients. Mylle *et al*. (2023) demonstrated a different force distribution strategy between Achilles tendinopathy patients and healthy controls. This altered strategy at the level of the triceps surae muscles determines altered force transmission to the AT, influencing the AT loading and, consequently, the AT strains. These outcomes underscore the complex nature of individual AT responses to load, emphasizing the importance of tailoring rehabilitation protocols to subject-specific characteristics, especially geometry and muscle forces. Despite that, the definition of an average ranking of rehabilitation exercises contributes to a general understanding of the diverse effects these exercises have on AT strains at a population level.

### 4.2 Strain distribution, peak location and peak strain values

Using subject-specific FE models for a large patient population demonstrated that strain distribution and peak strain location are influenced more by geometry than muscle forces. Regardless the type of exercise, the peak strain was consistently observed in the middle or upper mid-portion of the SOL sub-tendon, which is the most loaded sub-tendon. This aligns with findings indicating that tendinopathic tendons exhibit non-uniform tendon longitudinal morphology strain along their length under load, with the mid-portion undergoing larger strains (Nuri et al., 2018). Furthermore, the mid-portion is the location of the most pronounced pathological changes in tendon structure and composition in Achilles tendinopathy patients. Consequently, these changes in tendon thickness, fiber alignment, and collagen distribution can influence the mechanical behaviour of the tendon. The location of the peak strain in the mid-portion can also be explained by the fact that the least twisted geometry (corresponding to Type I twist described by Pękala et al. (2017)) was represented in this study. We decided to develop only this twisting geometry because it is the most common among individuals (Pękala et al., 2017) and, in our previous study, we demonstrated that the average strain in the mid-portion of the AT is not affected by the type of twist (Funaro et al., 2022). However, the twist influences the location of the peak strain, with Type I exhibiting the highest strain in the mid-portion of the tendon, for some exercises. Despite the peak strain consistently occurred in the mid-portion of the AT models, there was significant variability in the exact location of the peak among patients, as illustrated in **Figure 3**. These variations could be influenced by various factors, including the location of the swelling caused by tendinopathy or other attributes of the tendon’s geometry, including its shape, width, and length. Understanding this information could aid in predicting the likelihood of experiencing the highest strain in specific areas of the tendon for individuals with Achilles tendinopathy. This insight may also contribute to strategies for preventing or treating the condition. Therefore, additional research is necessary.

### 4.3 Limitations

There are some limitations of this study. The frictional contact between sub-tendons to account for the compromised sliding mechanism, which leads to more uniform deformations in the AT of Achilles tendinopathy patients (Couppé et al., 2020), was not used in our current FE model. It was not deemed essential to incorporate this feature into our current model to address our research questions. However, future development of FE models of Achilles tendinopathy patients will necessitate the incorporation of subject-specific frictional contact, to also take into account the alterations in non-uniform deformation within sub-tendons for estimation of the AT strains. In general, positive AT adaptation is not only dependent on strain magnitude but also time- and rate-dependent (Passini et al., 2021; T. Wang et al., 2015). However, the loading rate is a factor not accounted for in our model, given the decision to use a hyperelastic material as a constitutive model for the AT. The choice of hyperelastic material aligns with common practices in FE models of the AT (Handsfield et al., 2017; Hansen et al., 2017; Knaus & Blemker, 2021; Shim et al., 2019), assuming that tendons are in a ‘preconditioned state’ (Weiss *et al*., 1996). Given the application of load for a specific time point (time of peak total muscle force) during the execution of the exercise, the incorporation of a viscoelastic material was considered non-crucial. Subsequent development of FE AT models could explore the integration of viscoelastic material properties to assess the impact of loading rate on AT strains. Finally, while our study focused on tensile loads and the tensile strains of the AT, it is important to note that the mid-portion of the AT may also experience compressive loads (Pringels et al., 2022). However, as the rehabilitation exercises primarily involve tensile loading, we believed that it was reasonable to neglect the compressive load component in our current analysis. Further investigations with FE AT models could investigate compressive loads to provide a comprehensive understanding of AT strains during rehabilitation exercises.

### 4.4 Conclusions

In conclusion, we provided a ranking of various rehabilitation exercises in patients with Achilles tendinopathy based on the average strain in the mid-portion of the AT models. The analysis of individual rankings revealed noticeable variations in strain distribution patterns among participants. This finding points out the importance of tailored rehabilitation protocols which consider individual patient-specific morphological and material characteristics, and muscle forces. Furthermore, subject-specific FE models proved to be a valuable tool to explain the relationship between rehabilitation exercises and tendon strains. Our study highlights the significance of prioritizing not only subject-specific geometry but also subject-specific muscle forces when creating FE models aimed at the estimation of the AT strain in Achilles tendinopathy patients. The outcomes of this research present an important contribution to the future development of biomechanically informed rehabilitation protocols.

## 5. Ethics statement

The studies involving human participants were reviewed and approved by KU/UZ Leuven. The patients/participants provided their written informed consent to participate in this study.

## 7. Author Contributions

AF, VS, and BV contributed to conception and design of the study. AF performed volume reconstruction and segmentation, developed the model geometries, performed the free-form deformation, FE modelling and analysis, and statistical analysis. AF wrote the first draft of the manuscript. IM performed muscle force estimation. All authors contributed to manuscript revision, read, and approved the submitted version.

## Data Availability

All data produced in the present study are available upon reasonable request to the authors

## 9. Acknowledgments

The authors would like to thank the Research Council KU Leuven for providing financial support to this project.

## 9. Data Availability Statement

The data that support the findings of this study are available from the corresponding author 495 upon reasonable request.

## 10. Supplementary material

The Supplementary Material for this article can be found as additional tables attached to the submission.

