## Supplementary Material for "How subject-specific biomechanics influences tendon strains in Achilles tendinopathy patients: A finite element study"

### 1 Supplementary Tables

|  | Elastic modulus (MPa) |
| --- | --- |
| S01 | 503.29 |
| S02 | 319.26 |
| S03 | 774.84 |
| S04 | 959.25 |
| S05 | 406.05 |
| S06 | 793.95 |
| S07 | 647.1 |
| S08 | 754.35 |
| S09 | 645.12 |
| S10 | 478.48 |
| S11 | 628.57 |
| S12 | 709.56 |
| S13 | 320.6 |
| S14 | 386.98 |
| S15 | 810.74 |
| S16 | 1049.21 |
| S17 | 867.32 |
| S18 | 678.4 |
| S19 | 498.24 |
| S20 | 416.18 |
| S21 | 621.27 |
| average | 631.8457143 |

**Table 1**. Subject-specific elastic moduli and their average across 21 participants, identified by the code SX, where "S" denotes subject, and X represents the subject's assigned number.

- Bilateral heel rise

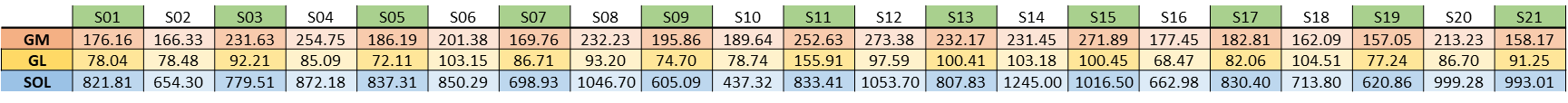

-
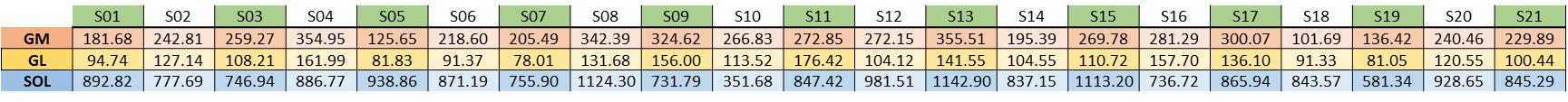
Bilateral heel drop
- Unilateral heel drop

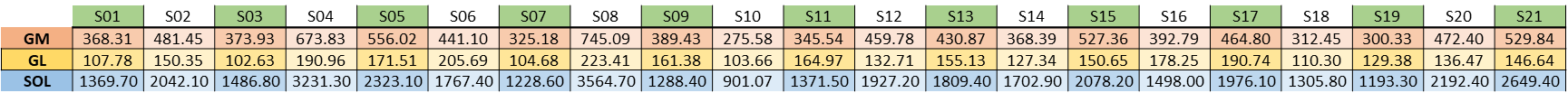

- Walking

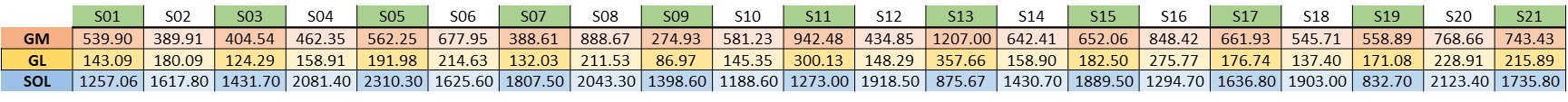

- Unilateral heel drop with flexed knee

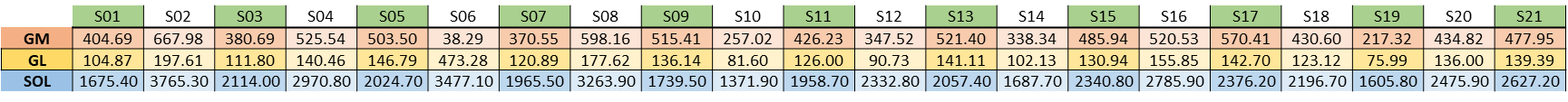

- Bilateral hopping

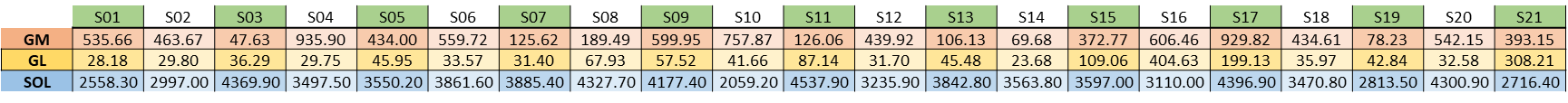

The tables illustrate the subject-specific muscle force values (expressed in Newton (N)) for the six rehabilitation exercises across the 21 participants, which serve as boundary conditions for loading the three sub-tendons (SOL, GM, and GL) of the FE models.
